# Psycho-social and behavioral trends in Type 2 Diabetes self-management amongst medically underserved patients during the COVID-19 pandemic in an mHealth intervention

**DOI:** 10.1101/2025.10.28.25338894

**Authors:** Elizabeth Campbell, Haomiao Jia, Andrea Cassells, TJ Lin, Jacqueline Cortez Lainez, Jonathan Tobin, Arlene Smaldone, Pooja Desai, George Hripcsak, Lena Mamykina

## Abstract

**Objective:** To examine behavioral and psychosocial aspects of Type 2 Diabetes Mellitus (T2DM) self-management among medically underserved adults during the COVID-19 pandemic.

**Materials and Methods:** Baseline data were analyzed from T2 Coach, a randomized controlled trial testing a behavioral mHealth intervention using a conversational agent to support T2DM self-management. Participants (n=279) were recruited from Federally Qualified Health Centers in underserved areas of the New York City metropolitan area (2020–2023). Measures included demographics, HbA1c, BMI, mobile device proficiency, and validated surveys assessing self-care adherence (SCI-R), self-efficacy, diabetes distress (PAID), and pandemic-related experiences (e.g., food insecurity, dietary and exercise changes). Multiple linear regression models examined associations between demographic, clinical, and COVID-19-related variables with SCI-R, self-efficacy, PAID, and HbA1c outcomes.

**Results:** Participants were primarily female (60.6%), Hispanic (63.4%), and foreign-born (65.9%), with substantial socioeconomic disadvantage (43.7% Medicaid enrollment; 42.7% food insecurity). Mean HbA1c was 10.1 (SD 1.7). Pandemic-related disruptions were common: 18.7% ate more, 34.5% exercised less, and ∼40% changed food purchasing habits. Regression models indicated that poorer self-reported health and dietary changes were linked to higher distress and lower self-efficacy. Increased exercise predicted better adherence (higher SCI-R), and women demonstrated stronger adherence than men. Older age was associated with lower HbA1c. Digital literacy was not associated with self-management outcomes.

**Discussion:** COVID-19 exacerbated challenges to diabetes self-management among underserved populations, particularly through lifestyle disruptions and psychosocial stressors.

**Conclusion:** Findings underscore the need for culturally responsive, technology-enabled interventions that address diet, exercise, and emotional well-being to improve diabetes outcomes during public health crises.

## BACKGROUND AND SIGNIFICANCE

Type 2 Diabetes Mellitus (T2DM) is a major public health issue in the United States. In recent decades, the US has seen a rise in chronic disease prevalence including T2DM. [1,2] The rising prevalence of overweight and obesity amongst adults in the United States greatly influences this trend as it is a known risk factor for many chronic conditions, particularly T2DM. [3]

Patients with T2DM are at risk for dangerous health complications and comorbidities, including cardiovascular disease, stroke, kidney disease, vision loss and blindness, and nerve damage. [4–6] T2DM is associated with an increased risk of various cancers, cognitive and functional disability, and hospitalization and mortality from infections. [6] Furthermore, rising T2DM prevalence carries significant financial costs associated with managing the disease, such as medical care utilization, medication, and lost productivity. The projected costs associated with T2DM in the United States exceed $300 billion annually. [7] T2DM is also a socially significant health issue. Racial and ethnic minorities, as well as low-income patients, have a higher prevalence of T2DM and associated comorbidities, in addition to facing limited access to healthcare services and education. [8,9]

Self-education, lifestyle modifications, and self-management support is critical to T2DM care. [10,11] The Association of Diabetes Care and Education Specialists identifies seven essential care behaviors: healthy coping (managing the emotional and psychological aspects of living with diabetes), healthy eating (making food choices that support blood glucose control), being active (engaging in regular physical activity), taking medication (using prescribed diabetes medications), monitoring (regularly checking blood glucose and other key health factors), reducing risks (taking preventative steps to avoid negative complications), and problem solving (working to prevent, detect, and respond to problems encountered in diabetes self-management). [12]

While T2DM self-management has always been a challenging practice, the COVID-19 pandemic ushered new issues that challenged patients’ self-management practices. The pandemic led to lifestyle changes, disruptions to routine care, increased psychological distress, reduced physical exercise, isolation, sleeping issues, and difficulty adhering to drug treatment. [13–17] In-person medical care and follow-up became less accessible during the COVID-19 pandemic, leading to a greater reliance on virtual resources for many T2DM patients. [18–21] Although the pandemic itself disproportionately impacted low-income and minority communities, [22,23] limited research has studied how the pandemic specifically affected T2DM self-management amongst medically underserved communities in the U.S. as well as individual factors that impact T2DM self-management outcomes for low-income and minority patients. [24,25]

Remote patient care and mHealth interventions have long played a significant role and have great potential in assisting T2DM patients with self-management activities. Such aid includes blood glucose monitoring, medication management, tracking diet and physical activity, providing educational resources, and facilitating community support. [26–28] However, adoption of mHealth technologies remains low among disadvantaged communities where low technology literacy presents a barrier to engaging with mHealth interventions. [29] The COVID-19 pandemic increased use and brought heightened awareness to the benefits of mHealth interventions for T2DM patients. However, there remains open questions as to whether higher digital literacy enabled individuals to persist with self-management during the COVID-19 pandemic.

## OBJECTIVE

The following cross-sectional study presents a secondary analysis of a dataset of baseline clinical and demographic characteristics for a randomized controlled trial (RCT) that tests the efficacy of a personalized behavioral mHealth intervention for individuals with Type 2 Diabetes. The trial used rolling recruitment and enrolled participants between 2020 and 2023, starting just prior to the COVID-19 pandemic, and continuing throughout its different phases. This study characterizes their T2DM self-management behavioral and psychosocial characteristics during the COVID-19 pandemic at the time they were randomized into the RCT. It aims to understand what patient-level factors are associated with higher or lower scores for self-management behavioral and psychosocial measures, and if there’s an association between patients’ T2DM self-management scores at baseline, pandemic severity, and pandemic duration. Given the increased reliance on virtual care during the pandemic, we also examine if digital literacy was associated with T2DM self-management and psychosocial outcomes. Our study aims to address the following research questions:

1. How can T2DM self-management be characterized during the COVID-19 pandemic amongst patients from medically underserved communities in a large urban area?
2. What patient-level clinical and demographic factors, along with variables related to COVID-19 exposure and experiences, are associated with T2DM self-management and psychosocial outcomes amongst medically underserved patients?
3. Was there an association between patients’ T2DM self-management and psychosocial outcomes at baseline and measures of pandemic severity and duration when patients entered the study?
4. Was there an association between digital literacy and T2DM self-management and psychosocial outcomes?

## MATERIALS AND METHODS

### Setting

The T2 Coach study is a two-arm RCT testing the efficacy of a personalized behavioral mHealth intervention for individuals with Type 2 Diabetes that provides personalized self-management coaching using combination of a chatbot delivered via text messaging and a mobile app for self-tracking. [30] T2 Coach uses Machine Learning (ML) with self-monitoring data to personalize coaching to each individual’s behavioral and glycemic patterns. [31] Participants for the trial were recruited from six Federally Qualified Health Centers (FQHCs) in medically underserved areas of the New York City metropolitan area. Recruitment began in January 2020 but was paused in March 2020 due to the onset of the COVID-19 pandemic. By August 2020, all components of the intervention had been switched to virtual and participant recruitment resumed. Participant recruitment was completed in September 2023.

### Study Population

The T2 Coach study utilized a two-arm RCT with 1:1 randomization at the participant level, with stratified randomization to balance by FQHC site, sex, and language. The total study population at baseline who provided consent, completed the baseline questionnaire, and were randomized in the study was 300 patients; 279 patients were recruited virtually after the COVID-19 pandemic’s onset and are the study population for this investigation.

The inclusion criteria for patients in the T2 Coach study were that participants were (a) a patient of the FQHC for ≥ 6 months and had a diagnosis of T2DM (b) had HbA1c ≥ 8.0, (c) were ages 18 to 65 years, (d) owned a smartphone, and (e) were proficient in either English or Spanish. After the onset of the pandemic, the protocol included an additional inclusion criterion: (h) able to download and install mobile apps, such as Zoom. Participants were excluded if they (a) were pregnant, (b) had a severe cognitive impairment (recorded in patient chart), (c) had other serious illnesses (e.g. cancer diagnosis with active treatment, advanced stage heart failure, dialysis, multiple sclerosis, advanced retinopathy, recorded in patient chart) (d) planned to leave the FHQC in the next 12 months, or e) had participated in the pilot study preceding T2 Coach. [32]

### Baseline Data Collection

At the baseline visit all subjects completed a demographic questionnaire, three diabetes questionnaires that assessed T2DM self-management and psychosocial measures, and a mobile device proficiency questionnaire (described below). Recruiters also conducted a medical records review using the site’s electronic medical records system. The demographic questionnaire was an investigator-developed instrument that collected self-reported data on several measures including age, race, ethnicity, gender, employment, income, and education. Current and past medical history and medications was collected from the patient chart and by study personnel as were clinical characteristics including patients’ HbA1c, fasting blood glucose, systolic blood pressure, diastolic blood pressure, lipids, weight, and BMI.

After the COVID-19 pandemic’s onset, an investigator_-_developed instrument was added to the baseline data collection protocol to assess the pandemic’s impact on numerous spheres of patients’ health and well-being. The instrument collected data on if patients had ever been tested for COVID-19, had ever tested positively for the illness, had ever been treated for COVID-19, and if they had received the vaccine. The survey also asked about patients’ self-reported health status, food insecurity, [33] the pandemic’s impact to self-management activities such as eating and exercise, experiences of stressful life changes, health promotion practices, anxiety, [34] sleep, [35] and depressive symptoms. [36] PROMIS measures for anxiety, sleep, and depressive symptoms are scored on a T-score metric, with a higher score corresponding to more of the concept being measured (e.g. a higher score on the depression questionnaire indicates that a patient is more depressed).

### Diabetes Questionnaires

Study personnel administered three surveys on patients’ self-care behaviors and psychosocial measures (Self Care Inventory-Revised Version (SCI-R), Self-Efficacy for Diabetes, and Problem Areas in Diabetes (PAID) self-management) at baseline, 6 months, and 12 months to monitor changes over the study’s duration. SCI-R, Self-Efficacy, and PAID data collected at baseline are used in this study.

The SCI-R is a validated questionnaire with fifteen questions that measures patients’ self-reported and self-perceived adherence to diabetes self-care recommendations. [37] SCI-R is designed for patients with Type 1 and Type 2 Diabetes. When administered to T2DM patients, patients are asked 12 out of 15 questions. Questions are scored on a Likert scale of 1-5, then summed and converted to a 0-100-point scale. A higher score indicates higher levels of self-care.

The Self-Efficacy for Diabetes scale is an eight-question questionnaire that surveys patients’ confidence in carrying out health promoting activities for T2DM self-management. Questions are scored on a Likert scale of 1-10, with a 1 meaning “not at all confident” and a 10 meaning “totally confident.” After survey completion, a respondent’s item scores are averaged into a final score ranging from 1-10, with a higher score indicating higher confidence in carrying out T2DM management activities. The scale has been validated and shows high internal consistency in English and Spanish language versions. [38]

The PAID is a 20-item questionnaire that surveys patients’ self-reported diabetes-related emotional distress. Questions are measured on a 0-4-point Likert scale, with a 0 indicating “not a problem” to a 4 indicating “a serious problem.” Patients’ responses are summed and converted to a 100-point scale for a final score, with higher scores indicating patients with higher levels of distress who need additional attention and resources. [39] PAID has been validated in English and Spanish versions which have demonstrated high reliability. [40]

### Digital Literacy Questionnaire

During baseline enrollment, study participants responded to a 16-question survey on self-perceived mobile device proficiency (MDPQ), which we use as a measure of digital literacy in this study. [41] Questions assessed participants’ confidence in a wide variety of mobile device uses, namely mobile device basics, communication, data and file storage, internet, calendar, entertainment, privacy, and trouble shooting and software management. MDPQ is scored on a scale of 0-40, with a higher score indicating greater technical literacy.

### COVID-19 Pandemic Severity Measures

Three variables were introduced into the model development process to contextualize the COVID-19 pandemic’s severity at the time that patients entered the T2 Coach Study: the 7-day averages in New York City for hospitalization counts, deaths by day, and case counts. Variable values were obtained for the day that baseline data was collected for each patient. This data is publicly available from the NYC Dept. of Health. [42] The year that patients entered the study was also included as a measure of pandemic duration.

### Statistical Analysis

Descriptive statistics for the demographic, clinical, and COVID-19 questionnaire variables collected at baseline were calculated to characterize the study population who was randomized to the intervention after the pandemic’s onset (n=279).

Four multiple linear regression models were developed to identify associations between demographic, clinical, and COVID-19 variables with the T2DM self-management behavioral and psychosocial outcomes of interest: SCI-R, Self-Efficacy, PAID, and HbA1c. Variables derived from baseline data collection, including the demographic questionnaire, MDPQ, BMI, and COVID-19 questionnaire were considered as independent variables for consideration in the model development process. Table 1 lists the dependent and independent variables used for model development in this study.

**Table 1.** Independent and Dependent Variables.

| Dependent Variables | Independent Variables |
| --- | --- |
| SCI-R<br>Self-Efficacy<br>PAID<br>HbA1c | *Year* (enrollment year)<br>*Site* (FQHC patient was enrolled at)<br>*COVID testing* (has the patient ever been tested for COVID-19)<br>*COVID results* (Has the patient ever tested positive for COVID-19)<br>*COVID treatment* (Has the patient ever been treated for COVID-19)<br>*Gender* (self-reported)<br>*Race/Ethnicity* (self-reported)<br>*Income* (Patient's self-reported combined annual family income)<br>*Health Insurance* (self-reported)<br>*Birth Location* (was a patient born in the US/ Puerto Rico or abroad)<br>*Education* (patient's self-reported highest level of education achieved)<br>*Marital Status* (self-reported)<br>*Language Spoken at Home* (self-reported)<br>*Food Insecure* (is a patient classified as food insecure based on Hunger Vital Sign)<br>*Changes to Ability to Buy Food* (within the past 6 months did your ability to buy food change because of the COVID pandemic?)<br>*Dietary Changes* (within the past 6 months did your diet change because of the COVID pandemic?)<br>*Exercise Changes* (within the past 6 months did your physical activity/ exercise change because of the COVID pandemic?) |
|  | *Starting Injectable Insulin* (Have you started using injectable insulin in the past 6 months)<br>*Bariatric Surgery* (Have you had bariatric surgery in the past 6 months?)<br>*Involvement in Health Promotion Programs* (Have you started any health promotion programs in the past 6 months?)<br>*Involvement in Mental Health Programs* (Have you started any mental or spiritual health programs in the past 6 months)<br>*Age* (Self-reported age in years)<br>*7-day Avg. Hospitalizations* (Average daily number of people hospitalized for COVID-19 in NYC in the 7 days before consent date)<br>*7-day Avg. Deaths* (Average daily number of people who died from COVID-19 in NYC in the 7 days before consent date)<br>*7-Day Avg. Cases* (Average daily number of people who tested positive for COVID-19 in NYC in the 7 days before consent date)<br>*MDPQ* (mobile device proficiency questionnaire score)<br>*BMI* (recorded by a healthcare professional)<br>*General Health* (patient's self-reported general health over the past 6 months, e.g. good, fair, poor)<br>*Stressful Life Events* (number of stressful events a patient had experienced in the past 6 months, e.g. divorce, loss of job)<br>*Anxiety T-Score* (PROMIS)<br>*Depression T-Score* (PROMIS)<br>*Sleep Disturbance T-Score* (PROMIS) |

Correlation coefficients were calculated for continuous independent variables with the dependent variables to check for linear association. Variables with a correlation coefficient > 0.15 were retained for bivariate regression analysis. Continuous variables with p <0.15 from the bivariate regression were included in the final multivariate analysis. ANOVA scores were calculated to test associations between categorical variables with the dependent variables. Categorical variables were retained for the multivariate analysis if p < 0.15. Four multiple linear regression models were fit for each dependent variable using the independent variables retained from the first analytical stage. The multiple R^2^ was used to evaluate each model’s goodness-of-fit.

In the PAID model, COVID treatment and COVID testing demonstrated multicollinearity and acted as suppressors for the outcome of interest. As a result, both variables were removed from the model. Variables capturing whether patients had begun a mental health program in the past six months and if they had begun a health promotion program in the past six months were correlated with HbA1c and SCI-R but were collinear. The variable capturing if patients had enrolled in a mental health promotion program was retained in the multivariate regression because a greater number of patients had enrolled in these.

## RESULTS

Table 2 presents the mean and standard deviation values for the dependent variables for the study population along with clinical, demographic, and technical literacy characteristics (n=279). Participants had a mean SCI-R score of 67.55, a mean PAID score of 30.04, a mean Self-Efficacy score of 5.83, and a mean HbA1c of 10.1.

**Table 2.** Clinical, Demographic, Self-Management, and Psychosocial Characteristics for Study Population (N (%))

| Indicator | Virtual<br>N= 279 |
| --- | --- |
| *Self-Management Behavioral and Psychosocial Measures* | Mean (SD) |
| *SCI-R* | 67.55 (12.62) |
| *PAID* | 30.04 (23.76) |
| *Self-Efficacy* | 5.83 (1.23) |
| *HbA1c* | 10.10 (1.70) |
| *Demographic and Clinical Characteristics* | N (%) |
| *Gender* |  |
| Male | 110 (39.4%) |
| Female | 169 (60.6%) |
| *Race/Ethnicity* |  |
| Black or African American | 80 (28.7%) |
| Hispanic | 177 (63.4%) |
| White | 11 (3.9%) |
| Multiple Race/Other Race | 11 (3.9%) |
| *Insurance* |  |
| Private | 33 (11.8%) |
| Medicare | 15 (5.4%) |
| Medicaid | 122 (43.7%) |
| None | 93 (33.3%) |
| Other | 16 (5.7%) |
| *Birth Location* |  |
| United States/ Puerto Rico | 95 (34.1%) |
| Another Country | 184 (65.9%) |
| *Language Spoken at Home* |  |
| English | 176 (63.1%) |
| Spanish | 100 (35.8%) |
| Other | 3 (1.1%) |
| *Education Level* |  |
| < High School | 85 (30.5%) |
| High School/GED | 86 (30.8%) |
| > High School | 108 (38.7%) |
| *Employment* |  |
| Yes | 174 (62.4%) |
| No | 105 (37.6%) |
| *Combined Family Income* |  |
| < \$10,000 | 97 (34.8%) |
| \$10,000- \$19,999 | 72 (25.8%) |
| \$20,000- \$39,999 | 64 (22.9%) |
| \$40,000- \$59,999 | 29 (10.4%) |
| >\$60,000 | 14 (5.0%) |
| Unknown | 3 (1.1%) |
| *Marital Status* |  |
| Married/Living with partner | 127 (45.5%) |
| Separated/ Divorced | 38 (13.6%) |
| Widowed | 11 (3.9%) |
| Never Married | 103 (36.9%) |
| *Age in Years (Mean (SD))* | 48.5 (9.2) |
| *BMI (Mean (SD))* | 33.2 (7.7) |
| *MDPQ (Mean (SD))* | 34.43 (6.00) |

More than half of the study population was female (60.6%), Hispanic (63.4%), and born outside of the United States (65.9%); approximately one quarter of participants were African American (28.7%). One third of participants did not have any health insurance and 43.7% were enrolled in Medicaid at baseline. Approximately two thirds of the study population completed a high school degree or less (61.3%) and were employed (62.4%); 83.5% had a combined family income of less than $40,000 per year. Nearly half of participants were married or lived with a partner (45.5%) while approximately one third of participants were never married (36.9%). The mean participant age at baseline was 48.5 years and the mean BMI was 33.2. The average MDPQ was 34.43.

Table 3 presents results from the COVID-19 questionnaire administered at baseline. Approximately half of participants enrolled in 2021 (50.9%) and 38.4% enrolled in 2022. Approximately two thirds of participants had been tested for COVID-19 multiple times (62.0%), and more than half of participants had reported never testing positive for COVID-19 (54.1%). One fifth of participants had tested positive for COVID-19 but received no treatment (21.9%) while 5% of participants had tested positive and received treatment. Participants had on average had at least one stressful life event (such as divorce, job loss, death of a loved one) in the past six months and 42.7% had experienced food insecurity. One in ten patients began injectable insulin in the last six months before study enrollment, 6.5% had joined a health promotion program, and 14.3% had joined a mental or spiritual health promotion program. On a scale of 1-5, with a 5 indicating poor health, participants reported a mean general health score of 3.3 for the past six months. Nearly 40% of participants had experienced some changes to their ability to buy food, 18.7% of participants reported eating more, and 34.5% reported exercising less in the last 6 months because of the COVID-19 pandemic. The mean PROMIS 4a Anxiety T-Score was 48.2, the mean PROMIS 4a Depression T-Score was 51.7, and the mean PROMIS 4a Sleep Disturbance T-Score was 46.15. The mean scores for sleep, depression, and anxiety were all within normal limits (defined as scores less than or equal to a T-score of 55) for patients in this study. [43]

**Table 3.** Summary Statistics for COVID-19 Questionnaire for Study Population (N (%))

| Measure | N (%) |
| --- | --- |
| *Baseline Enrollment Year* |  |
| 2020 | 17 (6.1%) |
| 2021 | 142 (50.9%) |
| 2022 | 107 (38.4%) |
| 2023 | 13 (4.7%) |
| *Site* |  |
| Site 1 | 24 (8.6%) |
| Site 2 | 39 (14.0%) |
| Site 3 | 50 (17.9%) |
| Site 4 | 59 (21.1%) |
| Site 5 | 57 (20.4%) |
| Site 6 | 50 (17.9%) |
| *Have you ever been tested for COVID-19?* |  |
| Never tested | 41 (14.7%) |
| Tested once | 63 (22.6%) |
| Tested more than once | 173 (62.0%) |
| Tested but number of times unknown | 2 (<1%) |
| *Have you ever tested positive for COVID-19?* |  |
| Never tested | 41 (14.7%) |
| Never positive | 151 (54.1%) |
| Tested positive | 84 (30.1%) |
| Testing results unknown | 3 (1.1%) |
| *Have you ever been treated for COVID-19?* |  |
| Never tested | 40 (14.3%) |
| Tested, never tested positive | 149 (53.4%) |
| Tested positive, no treatment | 61 (21.9%) |
| Tested positive, received treatment | 14 (5.0%) |
| *Food Insecure* |  |
| Yes | 119 (42.7%) |
| No | 160 (57.3%) |
| *Number of Stressful Life Events in the past 6 months (Mean(SD))* | 1.18 (0.88) |
| *Started injectable insulin in the last 6 months* |  |
| Yes | 28 (10.0%) |
| No | 251 (90.0%) |
| *Had bariatric surgery in the last 6 months* |  |
| Yes | 2 (<1%) |
| No | 277 (99.3%) |
| *Started any health promotion program in the last 6 months* |  |
| Yes | 18 (6.5%) |
| No | 258(92.4%) |
| Missing | 3 (1.1%) |
| *Started any mental or spiritual health program in the last 6 months* |  |
| Yes | 40 (14.3%) |
| No | 236 (84.6%) |
| Missing | 3 (1.1%) |
| *Self-Reported General Health (Mean (SD))* | 3.3 (0.93) |
| *Ability to Buy Food Changes* |  |
| Often True | 26 (9.4%) |
| Sometimes True | 83 (29.9%) |
| Never True | 166 (59.7%) |
| Don't know/ refused | 3 (1.1%) |
| *Changes to Diet* |  |
| Eating More | 52 (18.7%) |
| Eating Less | 59 (21.2%) |
| Eating Different Foods | 36 (12.9%) |
| No Change | 131 (47.1%) |
| *Changes to Exercise* |  |
| More Exercise | 29 (10.4%) |
| Less Exercise | 96 (34.5%) |
| Different Types of Exercise | 8 (2.9%) |
| No Change | 145 (52.2%) |
| *PROMIS 4a Anxiety T-Score (Mean (SD))* | 48.2 (10.13) |
| *PROMIS 4a Depression T-Score (Mean (SD))* | 51.7 (4.09) |
| *PROMIS 4a Sleep Disturbance T-Score (Mean (SD))* | 46.15 (8.50) |

Table 4 presents the multiple linear regression model results for the PAID, SCI-R, Self-Efficacy, and HbA1c models.

**Table 4.** Multiple Linear Regression Model Results for the PAID, SCI-R, Self-Efficacy, and HbA1c Models.

| **Measure** | **PAID**<br>***Beta (SE)*** | **SCI-R**<br>***Beta (SE)*** | **Self-Efficacy**<br>***Beta (SE)*** | **HbA1c**<br>***Beta (SE)*** |
| --- | --- | --- | --- | --- |
| *Intercept* | 14.13 (24.79) | 63.12 (4.60) | 7.39 (0.52) | 13.24 (0.94) |
| *Enrollment Year (ref: 2020)* |  |  |  |  |
| 2021 | -15.21 (6.94) * | -2.23 (3.55) |  | -0.89 (0.57) |
| 2022 | -9.49 (8.29) | -3.56 (3.55) |  | -0.96 (0.67) |
| 2023 | -4.86 (9.45) | -5.23 (4.79) |  | -1.25 (0.77) |
| *Site (Ref: Site 1)* |  |  |  |  |
| Site 2 | 1.81 (7.08) |  | -0.76 (3.14) | -0.81 (0.53) |
| Site 3 | 31.74 (6.96) *** |  | -0.32 (0.35) | -0.16 (0.46) |
| Site 4 | 4.98 (6.21) |  | 0.18 (0.31) | -0.13 (0.45) |
| Site 5 | 1.31 (5.71) |  | 0.29 (0.31) | -0.63 (0.41) |
| Site 6 | -9.22(6.02) |  | 0.06 (0.32) | -0.58 (0.43) |
| *Gender (Female vs Male)* |  | 3.46 (1.54) * |  |  |
| *Age in years* |  |  |  | -0.03 (0.01) ** |
| *Race/Ethnicity (ref: Black or African American)* |  |  |  |  |
| Hispanic | 2.87 (4.23) |  | 0.31 (0.23) |  |
| White | -4.02 (7.18) |  | -0.56 (0.41) |  |
| Other | -9.39 (7.08) |  | 0.54 (0.40) |  |
| *Insurance (ref: Private)* |  |  |  |  |
| Medicare | 2.55 (6.87) |  |  |  |
| Medicaid | 3.52 (4.20) |  |  |  |
| None | 0.98 (4.92) |  |  |  |
| Other | 6.45 (6.35) |  |  |  |
| *Birth Location (other country vs. US)* | -0.83 (3.73) |  |  |  |
| *Education (ref: <high school)* |  |  |  |  |
| High School/ GED | -4.88 (3.50) |  |  |  |
| More than High School | -6.27 (3.63) |  |  |  |
| *Marital Status (ref: Married/Living with a partner)* |  |  |  |  |
| Separated/ Divorced | 3.01 (3.76) |  |  |  |
| Widowed | -2.69 (6.54) |  |  |  |
| Never married | 2.79 (3.23) |  |  |  |
| *Language spoke at home (ref: English)* |  |  |  |  |
| Spanish | -5.00 (3.78) |  | -0.06 (0.21) |  |
| Other | -5.41 (3.78) |  | -1.16 (0.71) |  |
| *MDPQ Score* | -0.15 (0.25) |  |  |  |
| *Food Insecure (Yes vs. No)* | -5.32 (5.29) |  |  |  |
| *General Health Score* | 4.65 (1.40) ** |  | -0.28 (0.08) *** |  |
| *Changes to Ability to Buy Food (ref: Don't know/ refused)* |  |  |  |  |
| Sometimes true | -13.88 (13.19) |  |  |  |
| Never true | -17.85 (11.97) |  |  |  |
| Often true | -12.87 (13.53) |  |  |  |
| *Dietary Change (ref: No change)* |  |  |  |  |
| Eating Less | 4.29 (3.43) | -5.44 (2.05) ** | 0.26 (0.20) |  |
| Eating Different Foods | 12.39 (4.34) ** | -5.25 (2.55) * | 0.27 (0.25) |  |
| Eating more | 1.73 (3.61) | -5.10 (2.19) | 0.58 (0.21) ** |  |
| *Exercise Changes (ref: No change)* |  |  |  |  |
| Exercising Less |  | 2.14 (1.83) | 0.11 (0.17) |  |
| Different Exercises |  | 7.07 (4.64) | -0.75 (0.44) |  |
| Exercising more |  | 8.86 (2.58) *** | 0.40 (0.25) |  |
| *Injectable Insulin (No vs. Yes)* | -5.86 (4.17) |  |  |  |
| *Bariatric Surgery (No vs. yes)* | 8.82 (14.63) |  |  |  |
| *Anxiety T-Score* | 0.12 (0.17) |  |  |  |
| *Depression T-Score* | 0.42 (0.20) * |  | -0.02 (0.01) ** |  |
| *Started any mental or spiritual health program in the last 6 months (ref: yes)* |  |  |  |  |
| No |  | 2.84 (2.20) |  | -0.49 (0.31) |
| Missing |  | 12.07 (8.19) |  | 0.84 (1.10) |

\* 0.05 \*\* 0.001 \*\*\* 0.0001
Multiple R-Squared values for the PAID, SCI-R, Self-Efficacy, and HbA1c models were 0.42, 0.10, 0.19, and 0.09 respectively.

Four variables were associated with a statistically significant change to patients’ PAID score: site location, general health, dietary changes, and depression. MDPQ, which is a proxy for digital literacy, was not a significant predictor of PAID score. Patients’ self-described overall health of the past 6 months was scored on a scale of 1-5, with 1 indicating excellent health and 5 indicating poor health. Every 1-point increase on the scale corresponded to a 4.65-point increase to patients’ PAID score. Patients with poorer health encountered greater diabetes distress when managing their condition. Eating different foods over the past 6 months corresponded with a 12.39-point increase in PAID score. Patients with greater levels of depression reported higher levels of Diabetes-related distress. The multiple *R*^2^ was 0.42.

Women had significantly higher SCI-R scores, indicating greater adherence to T2DM self-management behavioral best practices. Patients who were eating less because of the pandemic had a 5.44-point decline in SCI-R scores compared to patients with no change. Patients who were eating different foods because of the pandemic had a 5.25-point decline in SCI-R scores compared to patients with no change. Patients who exercised more had an 8.86-point increase to SCI-R scores compared to patients with no change. The multiple *R*^2^ was 0.10.

Like the PAID score results, poorer general health over the past six months corresponded to lower self-efficacy in T2DM self-management. Every 1-point decrease in self-reported health corresponded to a 0.28-point reduction to patients’ Self-Efficacy score. Surprisingly, patients who were eating more because of the COVID-19 pandemic had a 0.58-point increase to Self-Efficacy scores compared to patients with no change to their diet. Patients reporting higher levels of depression also had significantly lower self-efficacy. The multiple *R*^2^ was 0.19.

Age was the only feature significantly associated with HbA1c. Older age was associated with lower HbA1c results. The multiple *R*^2^ was 0.09.

## DISCUSSION

The preceding study aimed to characterize behavioral and psychosocial trends in T2DM self-management amongst medically underserved communities in the New York City metropolitan area during the COVID-19 pandemic. Associations between patient-level clinical, technical literacy, and demographic factors, along with COVID-19 severity and duration measures with PAID, SCI-R, Self-Efficacy and HbA1c outcomes were investigated.

Measures of pandemic severity (average hospitalization counts at the time of randomization, death counts at the time of randomization, and case counts at the time of randomization) were not associated with psychosocial and behavioral trends in T2DM self-management. Year, which was a measure of pandemic duration, was generally not associated with T2DM self-management behavioral and psychosocial measures, except for patients who enrolled in the study in 2021 having lower rates of Diabetes-related distress. Individual interaction with the pandemic (COVID testing, COVID testing results, and COVID treatment) were also not associated with T2DM self-management psychosocial and behavioral outcomes. MDPQ, which is a proxy for digital literacy, was not a significant predictor for any behavioral and psychosocial outcome measured in this study.

Patients reported that the COVID-19 pandemic affected their general health and behaviors related to T2DM self-management, which in turn impacted psychosocial and behavioral outcomes. Diet and exercise are important in managing T2DM outcomes. The COVID-19 pandemic’s impacts on dietary and exercise changes were significantly associated with T2DM self-management behavioral and psychosocial measures. Patients who were eating different foods had significantly greater diabetes distress (higher PAID scores) and worse self-management behaviors (lower SCI-R scores) and eating less was significantly associated with lower SCI-R scores. Patients who were eating less and eating different foods may have had difficulty obtaining the usual foods that they use in T2DM self-management. Surprisingly, patients who ate more during the pandemic had higher self-efficacy scores compared to patients without dietary changes. Patients who began exercising more during the pandemic had significantly higher SCI-R scores. Finally, more severe depression was related to higher Diabetes-related distress and lower Self-Efficacy in managing T2DM, as was poorer self-reported general health.

Our study reflects findings from prior studies that identify the COVID-19 pandemic’s impact on T2DM self-care behaviors, mental stress, and overall well-being and characterizes T2DM self-management during the pandemic. [44–46] Most notably, it adds to the limited body of literature on the experiences of patients with T2DM from medically underserved communities in the United States during the COVID-19 pandemic. Our findings are similar to Zupa et al. [25] who investigated the pandemic’s impact on T2DM self-management practices among predominantly Latino patients at a FQHC in Detroit, a study population similar to our current investigation’s. Zupa et al. found that approximately half of T2DM patients reported less physical activity, two fifths had difficulty obtaining healthy food, and half experienced social isolation and stress. In our study, approximately two fifths of patients noted changes in their ability to buy food, one fifth reported eating more, and one third reported exercising less due to the COVID-19 pandemic; over 40% of participants were food insecure. Although participants in our study did not have high levels of anxiety, depression, or sleep disturbance, greater levels of depression negatively impacted Diabetes-related distress and self-efficacy at a statistically significant level. Like Vickery, et al., our findings also found that COVID-19 pandemic temporality was not directly associated with diabetes self-management behaviors or psychosocial outcomes and that behaviors related to T2DM self-management (diet, exercise, etc.) changed during the COVID-19 pandemic; however, our study did find a significant association between pandemic-induced changes to diet and exercise with psychosocial and behavioral measures of T2DM self-management. [24] Our study expands on Zupa et al. and Vickery et al. by investigating how pandemic severity measures and digital literacy were associated with T2DM behavioral and psychosocial outcomes.

### Strengths and Limitations

Our study’s findings are strengthened by the comprehensive number of independent variables including pandemic severity and duration measures, influences of the pandemic on factors that impact T2DM self-management outcomes such as diet and exercise, and numerous demographic and clinical factors. Data were collected across a three-year period during the COVID-19 pandemic, providing a broad picture of T2DM self-management behavioral and psychosocial trends throughout the pandemic. This allowed us to comprehensively assess T2DM self-management behavioral and psychosocial outcomes among a population of predominantly Latino and African American, immigrant, low-income patients in medically underserved communities; these patients were most vulnerable to the COVID-19 pandemic’s impacts on T2DM outcomes, [47,48] yet are underrepresented in scientific literature. Our findings are limited due to our study’s relatively small sample size and cross-sectional design. Despite baseline data collection occurring throughout the pandemic, longitudinal changes to T2DM self-management were not assessed for the study population. We expect our results are generalizable to T2DM patients from medically underserved communities in urban areas in the Northeast.

## CONCLUSION

Patients from medically underserved communities are most vulnerable to adverse T2DM outcomes. These patients were also the most affected by the COVID-19 pandemic’s changes to healthcare access and outcomes. Research into the COVID-19 pandemic’s impact to vulnerable patients’ T2DM self-management is urgent and critical, although this subject matter remains understudied. The preceding study examined the COVID-19 pandemic’s impact on four T2DM self-management behavioral and psychosocial outcomes using clinical, demographic, and technical literacy factors along with measures of the COVID-19 pandemic’s severity, duration, and impact to self-management behaviors. Future research may focus on exploring and characterizing patient dietary habits amongst vulnerable patients and how changes (or lack thereof) to these behaviors may have translated to the outcomes uncovered in this investigation. Our insights into behavioral and psychosocial trends in T2DM self-management during the COVID-19 pandemic amongst medically underserved communities offers insights into the factors that most influence these outcomes to inform education, policy, and healthcare interventions to mitigate negative impacts to T2DM self-management.

## Data Availability

The data that support the findings of this study are available from the corresponding author upon reasonable request.

## ACKNOWLEDGMENTS

The study team would like to thank the FQHCs that participated in this study: Open Door Family Medical Center (Ossining, NY), Open Door Family Medical Center (Port Chester, NY), Metropolitan Family Health Network (Jersey City, NJ), Morris Heights Health Center (The Bronx, NY), Family Physician Health Center at NYU Langone (Brooklyn, NY), and Bedford Stuyvesant Family Health Center (Brooklyn, NY).

## CONTRIBUTORSHIP STATEMENT

EC, AS, GH, JT, and LM contributed to conceptualization. EC, TL, and AC contributed to data curation. EC conducted formal analysis. EC and HJ contributed to methodology. EC contributed to writing (original draft). EC, AC, TL, JC, AS, PD, HJ, GH, JT, and LM reviewed and edited the manuscript. AC, PD, JC, and TL contributed to project administration. LM contributed to supervision and funding acquisition. All authors had final responsibility for the decision to submit for publication and agree to be accountable for all aspects of the work in ensuring that questions related to the accuracy or integrity of any part of the work are appropriately investigated and resolved.

